# Costing analysis to introduce a Contrast Enhanced Mammography service in place of existing breast MRI service for local staging of breast cancer

**DOI:** 10.1101/2022.08.18.22278958

**Authors:** Sarah L Savaridas, Huajie Jin

## Abstract

**Introduction:** Contrast-enhanced spectral mammography (CESM) is a functional imaging technique with comparable accuracy to MRI for loco-regional staging of breast cancer. This study assesses the cost impact of switching from CE-MRI to CESM for loco-regional staging of breast cancer from a public healthcare perspective.

**Methods:** The CE-MRI cost was obtained from NHS reference cost. The CESM cost was calculated using a bottom-up approach including use of the machine, pump injector, contrast, image storage and clinical staff’s time for reporting and cannulation. The cost of upgrading existing machines to CESM or purchasing new mammographic machines were obtained via national procurement. Other costs were obtained from local pharmacy, published unit cost data, or estimated based on surveys.

**Results:** For large health boards in Scotland (≥500 cancers diagnosed/annum), the cost savings of switching from CE-MRI to CESM range from £64,069 to £81,570. For small health boards (<500 cancers diagnosed/annum), the cost savings of switching from CE-MRI to CESM range from £6,453 to £23,953. The cost savings are most sensitive to the number of tests conducted per year, and whether the existing mammography machine can be upgraded to CESM or not.

**Conclusion:** Switching from CE-MRI to CESM for loco-regional staging of breast cancer is likely to be cost saving for both large and small health boards in Scotland. Further research is urgently needed to confirm the non-inferiority of CESM to CE-MRI as a locoregional staging technique. The input data of this analysis can be updated when such results become available.

**Highlights:**

- Switching from CE-MRI to CESM for locoregional staging is likely to be cost saving
- For large health boards, estimated annual savings range from £64,069 to £81,570
- For a small health boards, estimated annual savings range from £6,453 to £23,953
- Cost is driven by number of imaging studies and equipment upgrade vs replacement
- Research to confirm the non-inferiority of CESM for locoregional staging is needed

## Introduction

Contrast enhanced spectral mammography (CESM) is a functional imaging technique, which is becoming more widespread in clinical practice. It utilises a dual energy subtraction method following administration of intravenous contrast agent to produce mammographic images that demonstrate the vascularity of breast lesions.^1^ The benefits of functional breast imaging techniques are widely accepted with contrast-enhanced magnetic resonance imaging (CE-MRI), considered the gold-standard for loco-regional staging of breast cancer due to its high sensitivity.^2^ Use of MRI for pre-operative staging remains controversial due to concerns regarding false positive findings and the potential for this to result in inappropriate conversion from wide local excision to mastectomy.^3^ However, recent evidence suggests that increased mastectomy rates associated with MRI usage may be a result of the underlying nature of the disease being imaged, i.e. larger and multi-focal tumours, rather than the accuracy of MRI per se.^4^ Therefore, careful patient selection for pre-operative MRI is key. This is recognised in national guidance, which states that breast MRI is only indicated for loco-regional staging under the following circumstances:^5^

1. If breast conservation is being considered and sizing is uncertain on clinical evaluation and conventional imaging (mammography and ultrasound)
2. If breast-conserving surgery is being considered for invasive cancer with a lobular component
3. In mammographically occult tumours
4. Where there is suspicion of multifocal disease unconfirmed on conventional imaging
5. In the presence of malignant axillary node(s) with no primary tumour evident in the breast on conventional imaging
6. In Paget’s disease of the nipple if breast conservation is being considered. Unfortunately, due to pressures on clinical services it can be difficult to access MRI in a timely manner.^6^

An increasing body of evidence suggests that CESM may offer an alternative to MRI. In a recent meta-analysis, Xiang *et al* compare the diagnostic accuracy of CESM and MRI across thirteen studies. They conclude that CESM may be more effective than MRI, with a pooled sensitivity and specificity for CESM of 0.97 [CI:0.95-0.98] and 0.66 [0.59-0.71], compared with 0.97 [0.95-0.98] and 0.52 [0.46-0.58] for MRI.^7^ Although more limited, evidence of accuracy for loco-regional staging with CESM is also promising.^8–10^ This is reflected in national guidance, which states that ‘CESM has comparable accuracy to dynamic contrast-enhanced MRI for T-staging and assessing for multiple tumour foci’^5^

Emerging evidence from three studies of between 18 and 49 participants, demonstrates patient preference for CESM over MRI.^11–13^ This adds further weight to the argument that it may be appropriate to use CESM instead of MRI for loco-regional staging.

A further potential barrier to widespread introduction of CESM into clinical practice is the associated cost. Evidence from the American healthcare model suggests that CESM is more cost-effective than MRI in a private healthcare system.^14,15^ However this may not be directly applicable to a public healthcare system with an existing MRI service in place. This study aims to assess the cost impact of switching from CE-MRI to CESM for loco-regional staging for women with breast cancer, from the perspective of a public healthcare setting such as the NHS.

## Material and Methods

Per the Common Rule, ethical approval and informed patient consent were not required given that this is a costing analysis study with no direct patient contact or influence on patient care directly related to this work. Costings are calculated for ‘small’ health boards diagnosing less than 500 cancers per annum and ‘large’ health boards, diagnosing over 500 cancers per annum. As introducing a CESM service may require acquisition of new mammographic equipment or the upgrade of existing equipment, both scenarios are considered.

Radiology departments within each Scottish health board were contacted and asked to provide details on the number of breast MRI scans performed for loco-regional staging, within the health board per annum. Breast MRI scans performed for high-risk screening, implant integrity and monitoring response to neoadjuvant chemotherapy were excluded.

As there is no tariff available for CESM, costs were calculated using a bottom-up approach and included the following components:

- Use of the machine
- Contrast injector pump
- Contrast medium
- Image storage
- Clinical staff’s time for reporting and cannulation.

The cost of upgrading existing machines to CESM or purchasing new CESM machines and the cost of contrast injector pump were obtained via national procurement, NHS (National Health Service) National Services Scotland. The cost of iodinated contrast medium was obtained from the local hospital pharmacy.

To estimate radiologists’ reporting time surveys were conducted. Due to resource constraints of this project, snowball sampling was used to rapidly identify radiologists suitable for this project. This is a technique where identified stakeholders help to recruit additional stakeholders from among their colleagues or connections.^16^ With respect to CESM, consultant radiologists were asked to record the total number of studies they had reported to provide an indication of experience, and to estimate average time to report an individual study. Four UK sites with experience of CESM were contacted. Due to limited numbers of UK-based radiologists with experience of CESM, a further five European radiologists were also contacted. Regarding breast MRI, radiology departments providing breast imaging across Scotland were contacted. The survey was then cascaded to consultant radiologists routinely reporting full protocol CE-MRI studies for local staging. They were asked to record the total number of studies reported, and the estimated average reporting time. Results of these surveys was collated by the author, a consultant breast radiologist experienced in both CESM and MRI reporting. Health assistants time spent on cannulation was estimated based on expert opinion. The hourly rate for a radiologist and health assistant were obtained from the Personal Social Services Research Unit (PSSRU) 2020^17^.

The cost for CE-MRI, defined as ‘Magnetic Resonance Imaging Scan of One Area, with Pre- and Post-Contrast’, was obtained from the NHS reference cost, 2019/2020.^18^

## Results

With regards to MRI usage for loco-regional staging of breast cancer, responses were received from nine of thirteen Scottish health boards (see figure 1). Of these five provided exact figures. The remaining four provided estimates and were excluded from further analysis. For large health boards diagnosing over 500 breast cancers per annum an average of 212.5 MRIs were performed for local staging, ranging from 212 to 215. For smaller health boards diagnosing less than 500 breast cancers per annum an average of 70.3 MRIs were performed for the same indication, ranging from 42 to 123.

**Figure 1.**
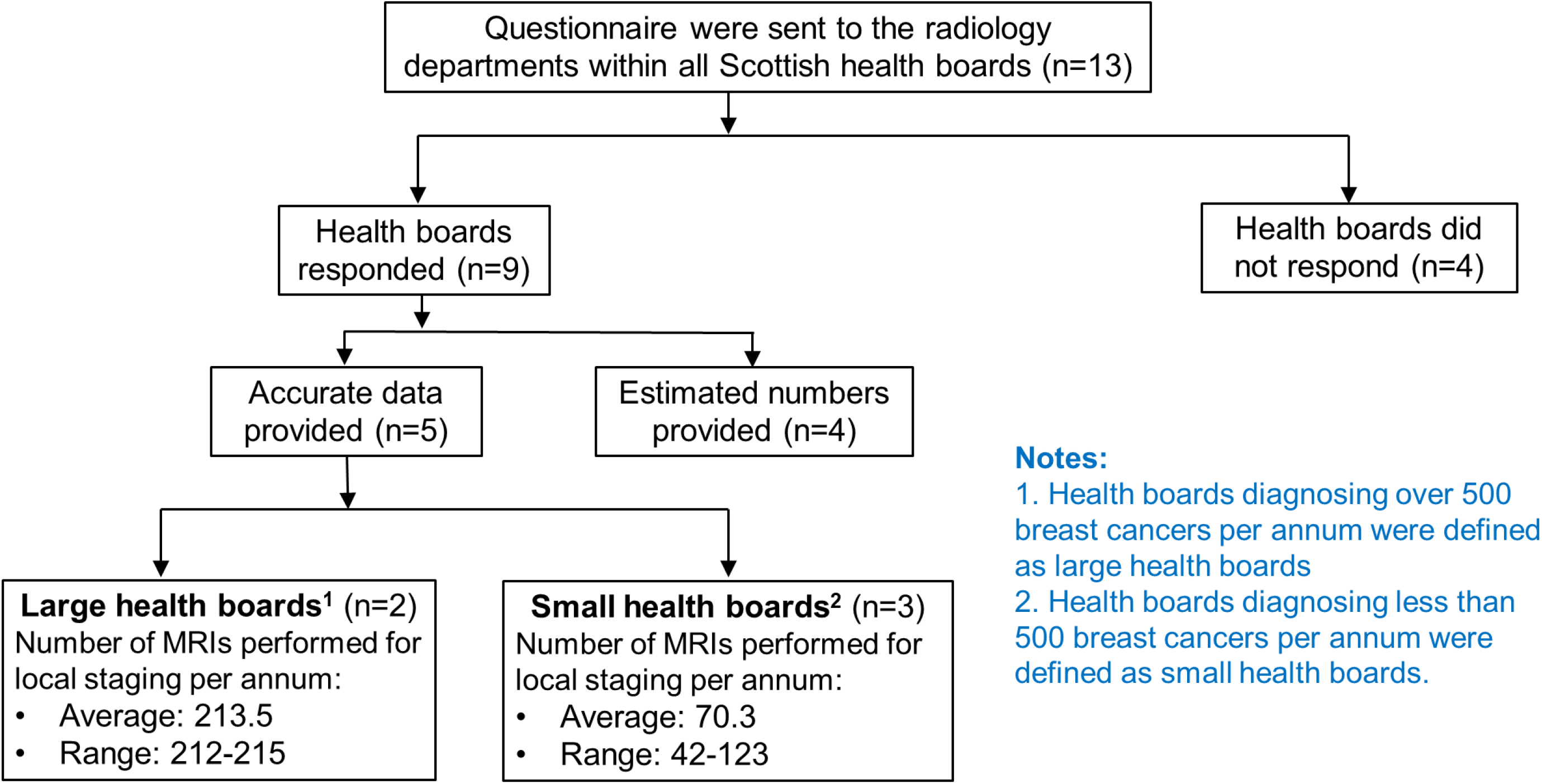
Flowchart of the survey process for estimating the number of breast MRI scans performed for local staging.

Of the 23 consultants responded to the survey regarding CESM-reading times (see figure 2). Of these eight had read over 100 CESM studies and four had read less than 50. The average estimated reporting time was 3.65 mins per study (range: 0.75 −10 mins). Twelve radiology consultants from seven Scottish health boards responded to the MRI reading-times survey. All had reported over 100 scans, many commenting they had reported thousands. Mean estimated reporting time was 20.63 mins (range: 10 – 45 mins). All input data for the costing analysis is summarised in table 1.

**Table 1.** Summary of input data.

| Input data | Base-case value | Min - Max value | Source |
| --- | --- | --- | --- |
| **Unit costs** |  |  |  |
| Purchase of new mammography machine | £217,000 | £217,000 - 285,000 | NHS national services Scotland |
| Upgrading existing mammography machine | £42,000 | Assumed fixed | NHS national services Scotland |
| Pump | £13,200 | £6,000 – 20,400 | Cost of reconditioned pump in Dundee |
| Contrast (per test) | £5.10 | Assumed fixed | Omnipaque 300 from the Forth Valley Royal Hospital pharmacy |
| Image storage | £2.98 | Assumed fixed | Uplifted from Public Health Scotland<sup>19</sup> |
| Hourly rate for radiologists | £73 | Assumed fixed | PSSRU 2020<sup>17</sup> |
| Hourly rate for healthcare assistant for cannulation | £34 | Assumed fixed | PSSRU 2020<sup>17</sup> |
| Cost per MRI | £425.18 | Assumed fixed | NHS reference cost 2019/20<sup>18</sup> |
| **Resource use and other input data** |  |  |  |
| Number of MRIs performed for local staging per large health board per year | 212.5 | 212-215 | Survey |
| Number of MRIs performed for local staging per small health board per year | 70.3 | 42-123 | Survey |
| Lifespan of a mammography machine | 10 years | Assumed fixed | Expert opinion |
| Lifespan of a pump | 10 years | Assumed fixed | Expert opinion |
| Reporting time for CESM per patient | 3.65 minutes | 0.75 – 10 mins | Survey |
| Time for healthcare assistant per cannulation | 5 minutes | Assumed fixed | Expert opinion |
| Reporting time for MRI per patient | 20.63 minutes | 12.5 - 37.5 mins | Survey |

**Figure 2.**
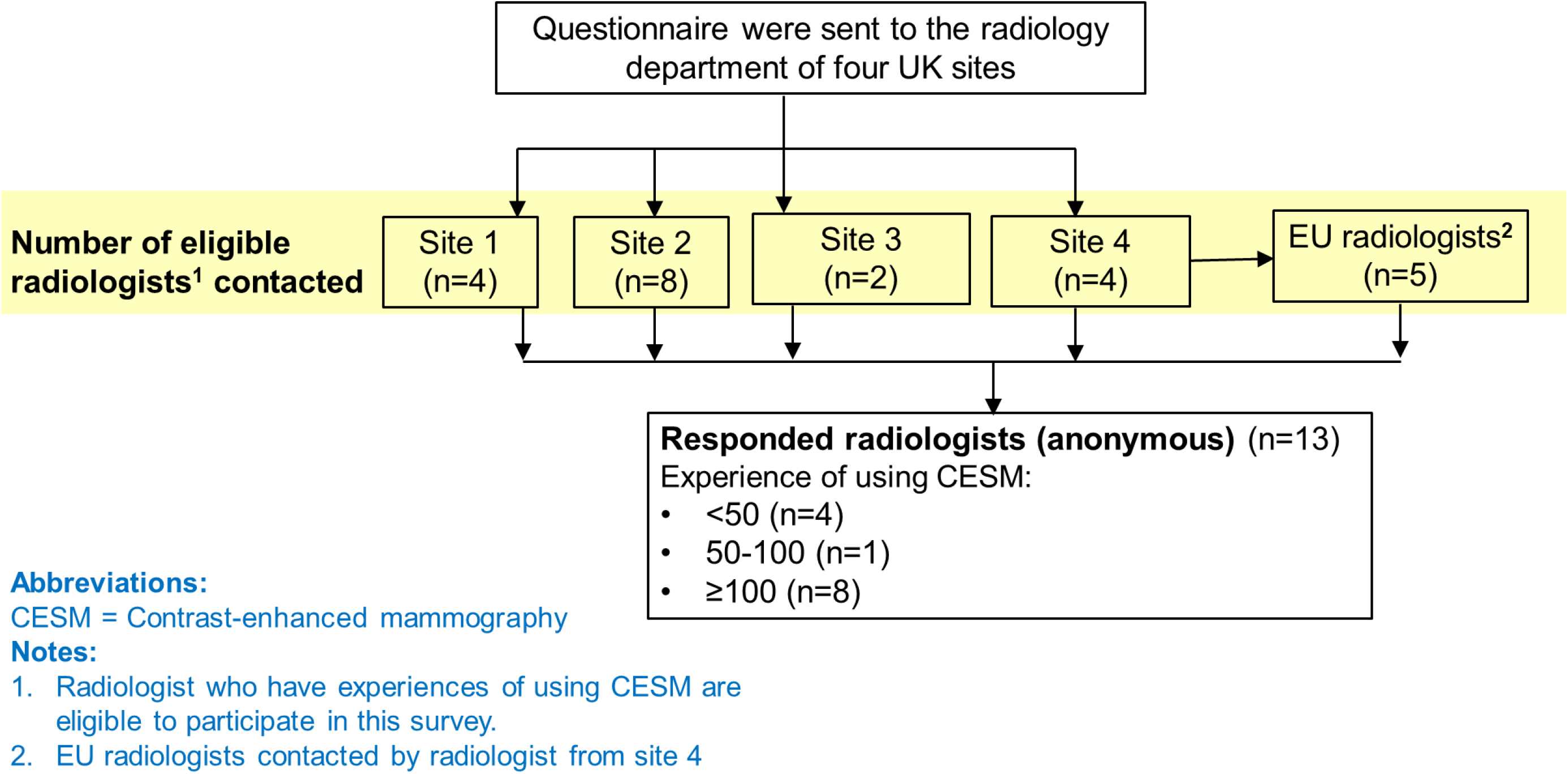
Flowchart of the survey process for estimating radiologists’ reporting time of CESM.

The CESM cost per patient for large and small health boards with detailed cost breakdown is reported in Table 2 and illustrated in Figure 3. The estimated cost of CE-MRI is £450.28 per study. For a small health board diagnosing less than 500 breast cancers per annum, the estimated cost of CESM per study is £84.45 (£47.30 – 204.68) if the existing mammography machine can be upgraded to CESM; and £333.39 (£189.57 – 783.25) if a new CESM machine needs to be purchased. For a large health board diagnosing over 500 breast cancers per annum, the cost of CESM per test is £41.32 (£32.68 −124.49) if the existing mammography machine can be upgraded to CESM; and £123.68 (£114.08 – 239.11) if a new CESM machine is required. Where it is possible to upgrade existing mammographic equipment, this equates to an average ten-fold cost saving for a large health board and a five-fold cost saving for smaller health boards; from £425.18 per MRI to £41.32 and £84.45 per CESM respectively. The savings are inevitably lower if the mammography equipment needs to be replaced, reducing to an average three-fold saving for large health boards. For smaller health boards replacing mammographic equipment, a cost-saving is not guaranteed due to the range in costings.

**Table 2.** Cost of CESM per patient for large health boards and small health boards

|  | Large health board |  | Small health board |  |
| --- | --- | --- | --- | --- |
|  | Purchase of new<br>mammography machine | Upgrade existing<br>mammography machine | Purchase of new<br>mammography machine | Upgrade existing<br>mammography machine |
| Machine/upgrade cost | £102.12 (£100.93-134.43) | £19.76 (£19.53-19.81) | £308.68 (£176.42-678.57) | £59.74 (£34.15-100.00) |
| Pump and injector | £6.21 (£1.33-81.60) | £6.21 (£1.33-81.60) | £9.36 (£1.33-81.60) | £9.36 (£1.33-81.60) |
| Contrast | £5.10 (assumed fixed) | £5.10 (assumed fixed) | £5.10 (assumed fixed) | £5.10 (assumed fixed) |
| Radiologists' time for reporting<br>(per patient) | £4.44 (£0.91-12.17) | £4.44 (£0.91-12.17) | £4.44 (£0.91-12.17) | £4.44 (£0.91-12.17) |
| Healthcare assistant's time<br>per cannulation | £2.83 (assumed fixed) | £2.83 (assumed fixed) | £2.83 (assumed fixed) | £2.83 (assumed fixed) |
| Image storage | £2.98 (assumed fixed) | £2.98 (assumed fixed) | £2.98 (assumed fixed) | £2.98 (assumed fixed) |
| **Total cost per CESM study** | **£123.68** (£114.08 – 239.11) | **£41.32** (£32.68 – 124.49) | **£333.39** (£189.57 – 783.25) | **£84.45** (£47.30-204.68) |

**Figure 3.**
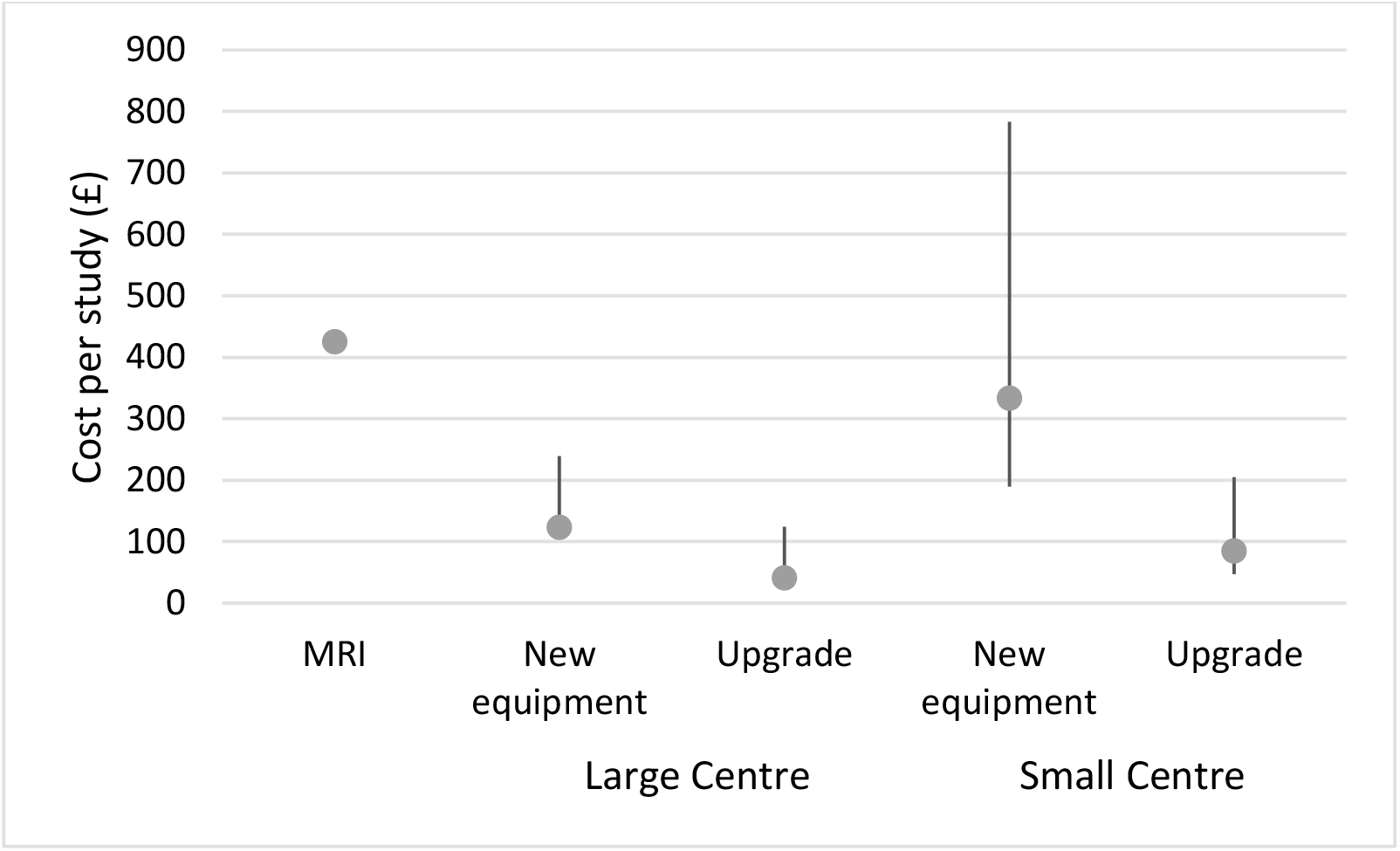
Cost of CESM per patient for large health boards and small health boards with respect to MRI.

Potential annual savings are presented in table 3, based on average number of MRIs performed for local staging per annum. For a large health-board this equates to average annual savings of £81,570 and £64,069 for equipment upgrade or replacement respectively. For a small health board, the average annual savings drop to £23,953 and £6,453 with the potential for costs to be incurred for equipment replacement, range: -£25,172 to £16,563.

**Table 3.** Annual Cost savings of CESM for large health boards and small health boards with respect to MRI

|  | Large health board<br>(212.5 studies per annum) |  | Small health board<br>(70.3 studies per annum) |  |
| --- | --- | --- | --- | --- |
|  | Annual cost (£) | Annual saving (£) | Annual cost (£) | Annual saving (£) |
| **MRI** | 90,351 | - | 29,890 | - |
| **CESM**<br>(New machine) | **26,282**<br>(24,242 - 50,811) | **64,069**<br>(39,540 – 66,109) | **23,437**<br>(13,327 – 55,062) | **6,453**<br>(-25,172 – 16,563) |
| **CESM**<br>(upgrade) | **8,781**<br>(6,945 - 26,454) | **81,570**<br>(63,897 – 83,406) | **5,937**<br>(3,325 – 14,389) | **23,953**<br>(15,501 – 15,501) |

## Discussion

CESM has the potential to offer an alternative to breast MRI for certain situations, including loco-regional staging. As a technique it is more time-efficient and often preferred by patients.^11–13^ Furthermore, switching to CESM would reduce the pressure on MRI services for other imaging studies.

Widespread uptake of CESM within the NHS may be limited by the perceived cost of introducing the service. To our knowledge this is the first study to do a detailed costing analysis, specific to public healthcare.

As expected, cost of CESM is driven by the number of tests performed. The introduction of a CESM service is highly likely to be cost saving for large health board. With potential annual savings in excess of £80,000 per annum if existing equipment can be upgraded, and £64,000 if a new mammography unit is required. Whilst it may not always be cost saving for a small health-board to purchase a new mammography machine for the sole purpose of performing CESM, this would be a highly unusual clinical scenario. If existing equipment could be upgraded, we suggest that the introduction of a CESM service would also be cost saving for small health board. Our findings are consistent with existing evidence based on American data. Patel *et al* conclude that in the context of screening, if CESM was to replace MRI, savings would be approximately $750 per examination and as much as $1.1 billion annually.^14^ In a conference abstract, Rane *et al* conclude that if CESM was used as an alternative to MRI for pre-operative local staging the net savings would be $36,530,278 with a total saving per cancer diagnosis of $1,282.^15^

This presents an unusual but very positive scenario, whereby the health service can save money whilst providing a service with equivalent accuracy and preferred by the majority of patients.

There are several limitations of the economic analyses presented here. Non-inferiority of CESM compared to MRI for loco-regional staging was assumed based small studies^9,20^ and meta-analysis data of diagnostic accuracy^7,21,22^ derived from primary research studies conducted in Central and North America, mainland Europe, Egypt, Taiwan and China. There is currently a lack of UK-specific clinical evidence directly comparing the accuracy of CESM with MRI for loco-regional staging of breast cancer. Therefore, it was only possible to compare the short-term costs of using CESM with MRI. In addition, this study cannot account for differing work-flow, for example the number of additional biopsies generated by the differing techniques. When future clinical evidence become available in the UK, we recommend our analysis be updated to incorporate any accuracy difference between the imaging techniques, and subsequent long-term cost and health impacts of using CESM and MRI. It is another limitation of this study that, when calculating the unit cost of using CESM machine, we only consider the use of CESM for loco-regional staging according to UK national guidelines. In practice, CESM may be used for broader purposes, such as loco-regional staging for women with dense breasts, monitoring response to neoadjuvant chemotherapy, high risk breast screening and screening assessment.^23^ The unit cost of CESM will reduce significantly as the number of required tests increases. Therefore, if all indications were considered, it is likely CESM would be cost saving even for a small health board requiring a new mammography machine. There are also logistic considerations which may impede the introduction of this service, as CESM imaging would be conducted within the breast unit by radiographers trained in mammography. Although the introduction of a service would relieve the pressure on the MRI service elsewhere within the hospital, it may initially increase pressure on breast imaging services. Finally, the number of radiologists contacted regarding reporting times was limited. It is anticipated that more accurate reporting times data will be acquired as part of future prospective imaging studies.

In conclusion the introduction of a CESM service to replace CE-MRI for loco-regional staging of breast cancer is likely to be cost saving for both large and small health boards. The unit cost of CESM would reduce significantly as the number of required tests increases. Further research, such as a prospective clinical trial, is needed to confirm the non-inferiority of CESM to CE-MRI as a loco-regional staging technique. With publication of such results, the input data of these analysis can be updated to assess the long-term cost and health implications of switching from CE-MRI to CESM.

## Data Availability

All data produced in the present work are contained in the manuscript

## Abbreviations

CESM: : Contrast enhanced spectral mammography
CE-MRI: : Contrast enhanced magnetic resonance imaging
PSSRU: : Personal Social Services Research Unit
NHS: : National Health Service

